# A Systematic Review of Randomised Controlled Trials on the Effectiveness of Ecotherapy Interventions for Treating Mental Disorders

**DOI:** 10.1101/2020.09.25.20201525

**Authors:** T. Williams, G.C. Barnwell, D.J. Stein

## Abstract

**Background:** Systematic reviews on ecotherapy interventions (i.e. environmental therapies and animal-assisted therapies) in the general population have demonstrated promising findings. However, there is a need for systematic assessment of the evidence for the use of these interventions in people living with mental disorders. Hence, we conducted a systematic review of randomised control trials (RCTs) on exposure to ecotherapy interventions (i.e. environmental therapy, animal-assisted therapy, wilderness therapy) for individuals with mental disorders.

**Methods:** The search was completed in September 2019 and comprised: Science Direct, PubMed Central, EBSCOHOST (via Academic Search Premier), the Cochrane Library, and Google Scholar. The primary outcomes that were assessed include: treatment efficacy (on the CGI-I or similar), symptom severity, and/or dropout rates. Secondary outcome measures assessed included self-efficacy, perceived control, hope, quality of life, life skills, and coping.

**Results:** A total of 2415 reports were identified, of which 94 were assessed for eligibility. Eight environmental therapy trials (i.e. gardening, forest therapy, horticultural therapy, nature adventure rehabilitation, and adventure-based therapy) and ten animal-assisted therapy trials (i.e. dogs, horses or dolphins) were included in the review. Risk of bias assessment was conducted, and qualitative analysis performed to describe the efficacy of the RCTs. The findings suggest that environmental therapies or animal-assisted therapies are efficacious in reducing symptoms of anxiety, depressive, substance-related and addictive, schizophrenia-spectrum and trauma- and stress-related disorders.

**Conclusion:** There is suggestive, but not conclusive, evidence for the efficacy of some ecotherapy interventions in mental disorders. In particular, there are: four environmental therapy trials and five animal-assisted therapy trails for depression and/or anxiety; one environmental therapy trial for post-traumatic stress disorder; three environmental therapy and animal assisted therapy trials for general psychiatric disorders; one environmental therapy trial and two animal assisted therapy trials for substance use disorders; and, one environmental therapy and two animal-assisted trials for people living with schizophrenia spectrum disorder. The use of standard reporting guidelines may improve evidence quality of future ecotherapy RCTs, and provide a foundation meta-analysis of the evidence.

## 1 Introduction

Mental disorders, excluding neurological disorders, affect around 1 billion people globally and account for 7% of the global burden of disease, causing premature death and ill health and increased risk of high medical and societal costs (Rehm &Shield, 2019; The World Health Organisation, 2001, 2013). Evidence of the burden of mental disorder has helped focus attention on the social and environmental determinants of mental disorders, in the hope that these might lead to targeted interventions to address these conditions (Lancet Commission on Mental Disorders and Sustainable Development).

Studies on the impacts of environments on mental health grew rapidly in the 1960s alongside growing environmental concerns (Steg, et al., 2018). There is growing evidence that lack of green spaces and presence of pollution are associated with an increased risk of depression and anxiety (Nutsford, et al., 2013; Power, et al., 2015; Rautio, et al., 2019; Tomita, 2017). Positive associations have been found between poverty indicators, such as food insecurity and poor housing conditions, with common mental disorders (Lund, et al. 2010). It has also been suggested that pollution may be a possible risk factor in schizophrenia (Attademo, et al., 2017; Gao, et al. 2017). While it is well accepted that poor environmental conditions are bad for mental health, exposure to the natural environment may also assist in recovery.

Anecdotal evidence for the benefits of nature on health extends at least as far back as the ancient Greek authors. However, in 1984, environmental psychologist Robert Ulrich published a paper called View Through a Window May Influence Recovery from Surgery.

The paper shared results of the first comparison ecotherapy study (N=46) where half of the post-surgery patients were assigned to a room with their windows facing a brick wall, while the other half looked out over a garden. Ulrich found that those with a view of trees had shorter postoperative stays compared to their counterparts. Numerous studies have since emerged to understand the links that exist between exposure to nature and patient recovery. Yet, it is only recently that ecotherapy interventions have been developed and deployed by clinicians as primary or complementary interventions to manage and treat mental illness (Buzzell &Chalquist 2009; Jordan &Hinds 2016). Ecotherapy has become an emerging clinical modality and an umbrella term for techniques and practices that draw from the natural world’s therapeutic benefits. Modalities include environmental therapies (e.g. horticultural therapy, gardening, forest bathing) and animal-assisted therapy.

Systematic reviews have been conducted on environmental therapies and animal-assisted therapies in healthy individuals (Kamioka, et al., 2014; Kondo, et al., 2018; Lee, et al., 2017; Wen, et al., 2019; Roberts, et al., 2019). These reviews raise concerns over the methodological rigour of the evidence-base (Kamioka, et al., 2014; Kondo, et al., 2018; Wen, et al., 2019; Roberts, et al., 2019). For instance, a systematic review on forest bathing, as an environmental therapy, found significant improvements in anxiety and depressive mood, and general enhancements in emotional states in healthy individuals (Wen, et al., 2019). Similarly, forest bathing has been shown to improve adult depressive features in the general population (Lee, et al., 2017). Although, Roberts et al. (2019) found contradictory evidence after they reviewed 33 studies on short-term exposure to the natural environment on depressive mood in healthy individuals, finding small variation in effect size. The limited number of high-quality studies on ecotherapies also creates barriers for issuing detailed clinical guidance (Oh, et al., 2017). That said, ‘Green prescriptions’ are being considered more widely as a cost-effective complementary intervention (Shanahan, et al. 2019). It would be timely, therefore, to undertake a systematic review of randomised controlled ecotherapy trials for the treatment of mental disorders. Therefore, we undertake a review of such trials.

## 2. Aims and Objectives

### 2.1 Aim

To systematically review randomised control trials on ecotherapy interventions for the treatment of psychiatric disorders.

### 2.2 Objectives

#### Objective 1

To describe the efficacy of randomised control trials on ecotherapy interventions for the treatment of psychiatric disorders.

#### Objective 2

To assess the quality of evidence (i.e. CONSORT) of randomised control trials on ecotherapy interventions for the treatment of psychiatric disorders;

## 3. Methods

This section outlines the methodology employed by the systematic review.

### 3.1 Data Collection and Search Strategy

Science Direct, PubMed Central, EBSCOHOST (via Academic Search Premier), the Cochrane Library, and Google Scholar were all searched for therapies as defined by nature (i.e. environmental therapy and animal-assisted therapy) published up until November 2015, including reference lists of related articles. A comprehensive review was concluded in September 2019 to update the previous search.

#### 3.1.1 Search Terms

The following search terms were used: Mental Disorders [MeSH] OR ‘Psychiatric illness’ OR ‘Psychiatric diseases’ OR ‘Diagnosis, Psychiatric’ OR ‘Psychiatric Diagnosis’ OR ‘Mentally Ill Persons’ AND Nature [MeSH] OR ‘Plant’ OR ‘Ornamental Plants’ OR Animal [MeSH] OR ‘Animal’ OR ‘Animalia’ OR ‘Metazoa’ AND Randomized Controlled Trials [MeSH] OR ‘Clinical Trials, Randomized’ OR ‘Controlled Clinical Trials, Randomized’ OR ‘Trials, Randomized Clinical’. In addition to these terms the following terms were used by previous studies and were added to the search strategy: Animal-Assisted Therapy [MeSH] OR ‘Animal Facilitated Therapy’ OR ‘Pet Facilitated Therapy’ OR ‘Pet Therapy’ OR Equine-Assisted Therapy [MeSH] OR ‘Equine-Assisted Psychotherapy’ OR ‘Hippotherapy’ OR ‘Horseback Riding Therapy’ OR ‘Recreational Horseback Riding Therapy’ OR ‘Green Care’ OR ‘Green Space’ OR ‘Horticultural Therapy’ OR ‘Wilderness Therapy’ OR ‘Forest Therapy’ OR ‘Outdoor Gardening’ OR ‘Outdoor Adventure Therapy’ OR ‘Ecotherapy’.

#### 3.1.1 Eligibility Criteria

We included all randomised controlled trials of therapies comprising of environmental therapy and animal-assisted therapy compared with a control group or alternative intervention for the treatment of psychiatric disorders. No restrictions were placed on language, gender, publication date and setting. However, participants between the age of 18 years and older with a psychiatric disorder defined by the Diagnostic Statistical Manual for Mental Disorders (DSM) (i.e. Axis I and II) or other specified measure were included. Studies focusing on behavioural, cognitive and neurocognitive disorders were excluded as well as general medical conditions such as cancer or pain disorders. Preventative studies were also excluded.

### 3.2 Description of Search Results

A total of 2415 study reports were found through the search process (Science Direct 540, PubMed Central 98, EbscoHost 329, the Cochrane Library 31, and Google Scholar 1417). The title and abstract of each study were screened for eligibility, and seventy studies initially seemed relevant. After further inspection, we excluded 76 of these studies. These studies were excluded because the disorders were not psychiatric disorders defined within our inclusion criteria. Eighteen RCTs met study inclusion and were included in the review. Figure 1 presents the Prisma diagram.

**Figure 1.**
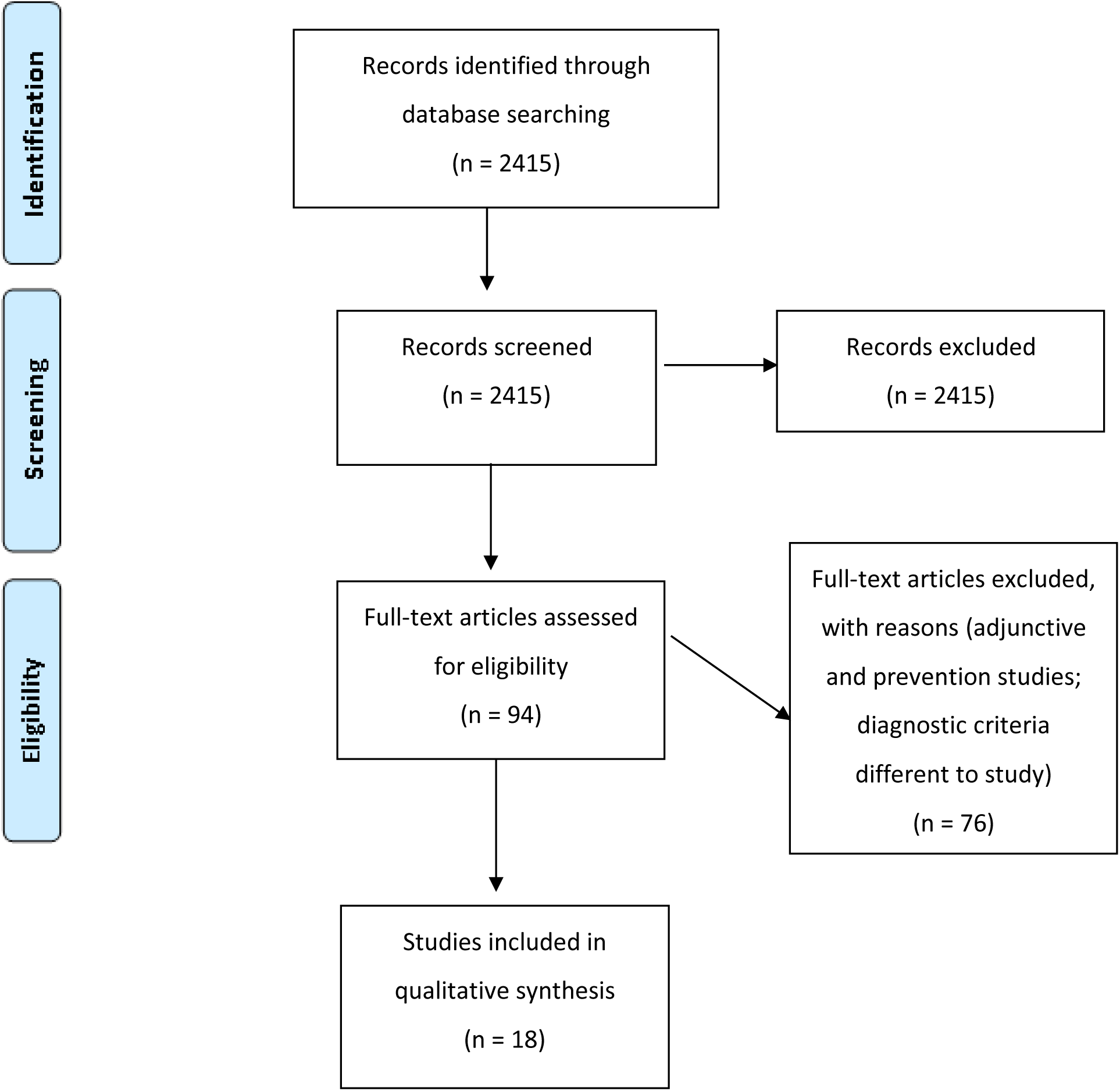
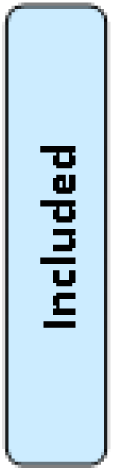
Prisma diagram - Study Eligibility.

### 3.3 Data Extraction

Data extraction included the following data items (See *Table 1: Study Characteristics*): 1) study ID (author, year); 2) health outcomes; 3) funding; 4) country/setting; 5) intervention; 6) comparison group; 7) weeks/days; 8) total sample; 9) mean age of sample/range; 10) mental health measures; 11) drop-outs; 12) loss to follow up. Furthermore, CONSORT 2010 Checklist (i.e. standards for information to include in the reporting of randomised control trials) items were used for extraction for the assessment of the quality of evidence.

**Table 1:**
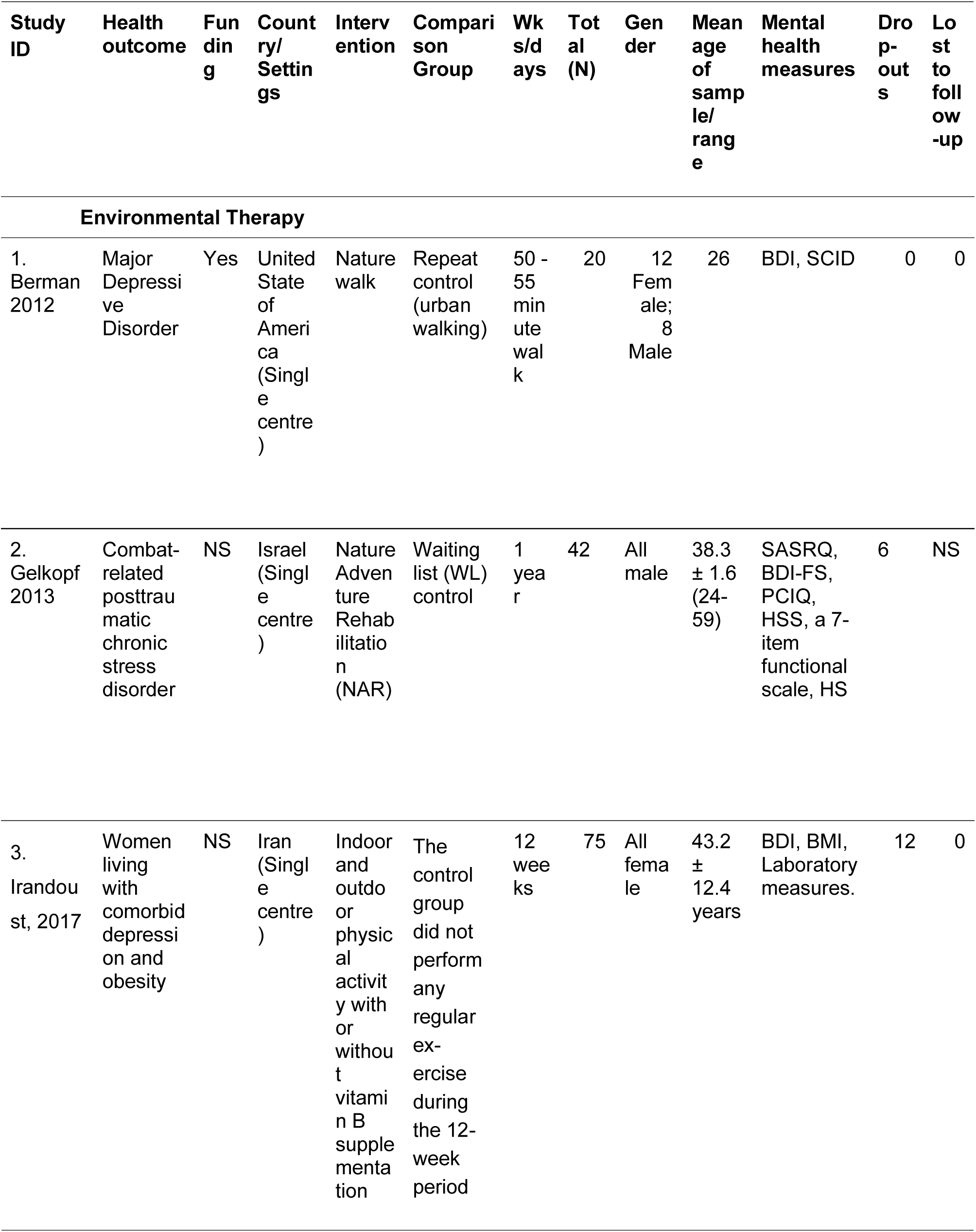
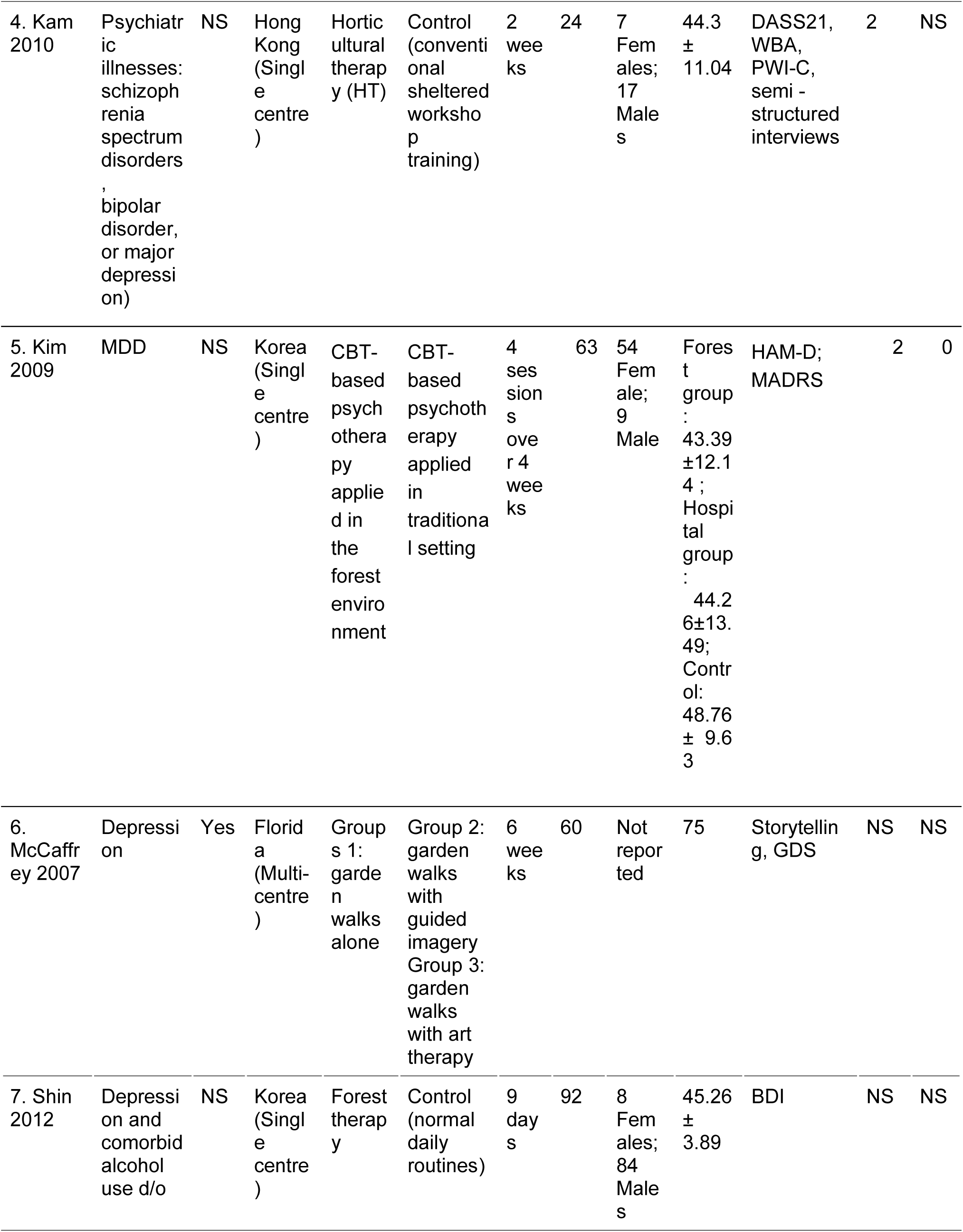
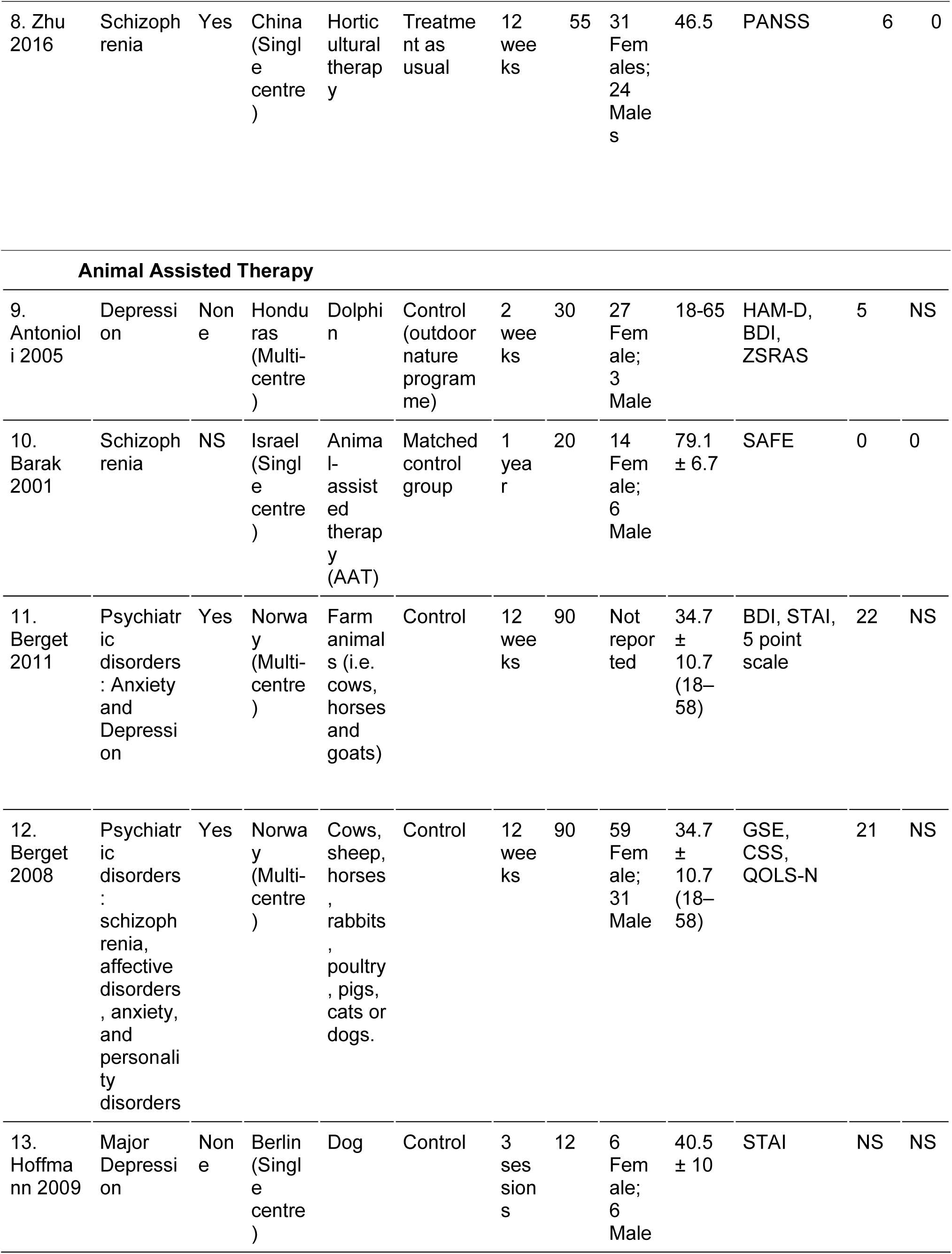
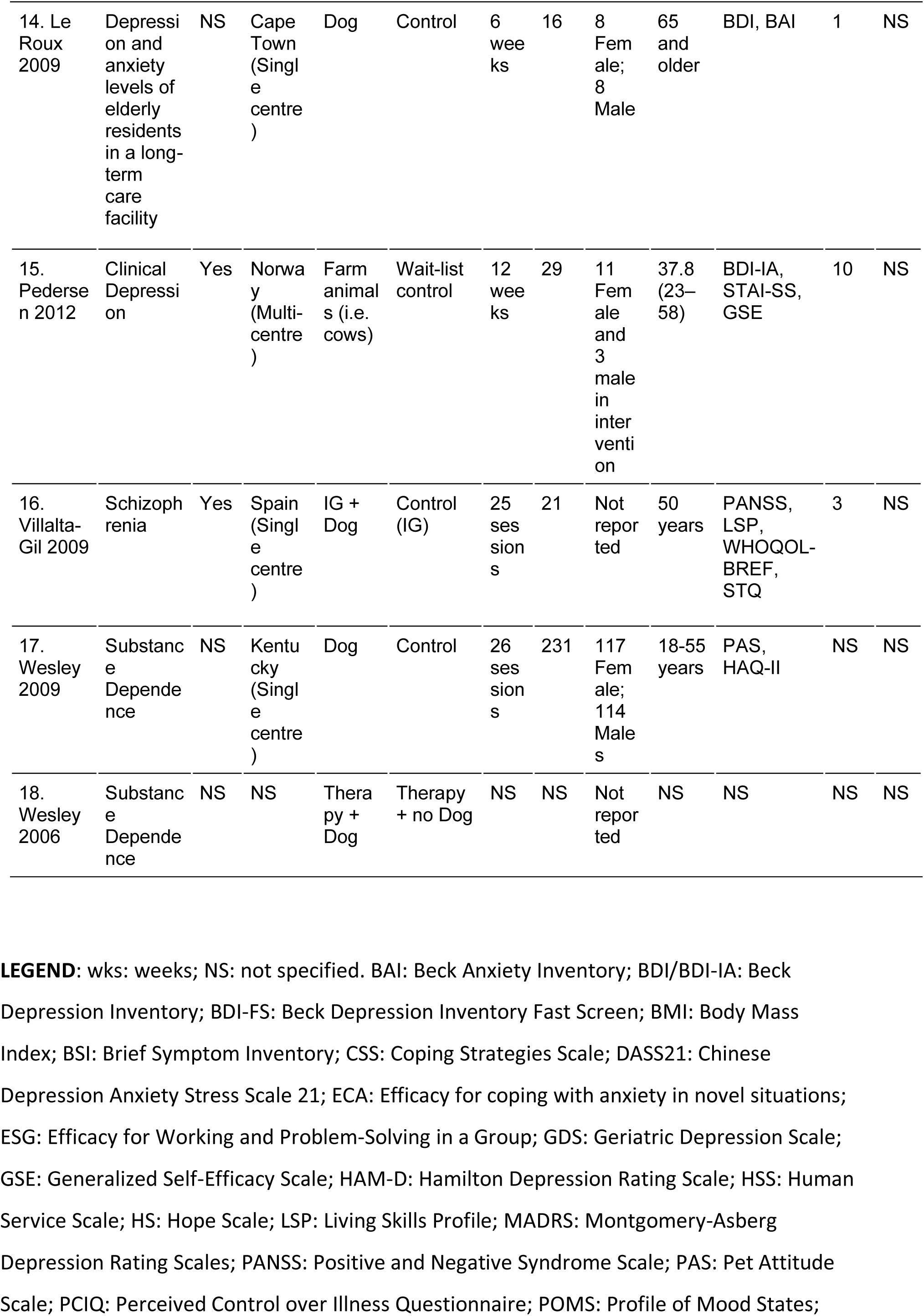
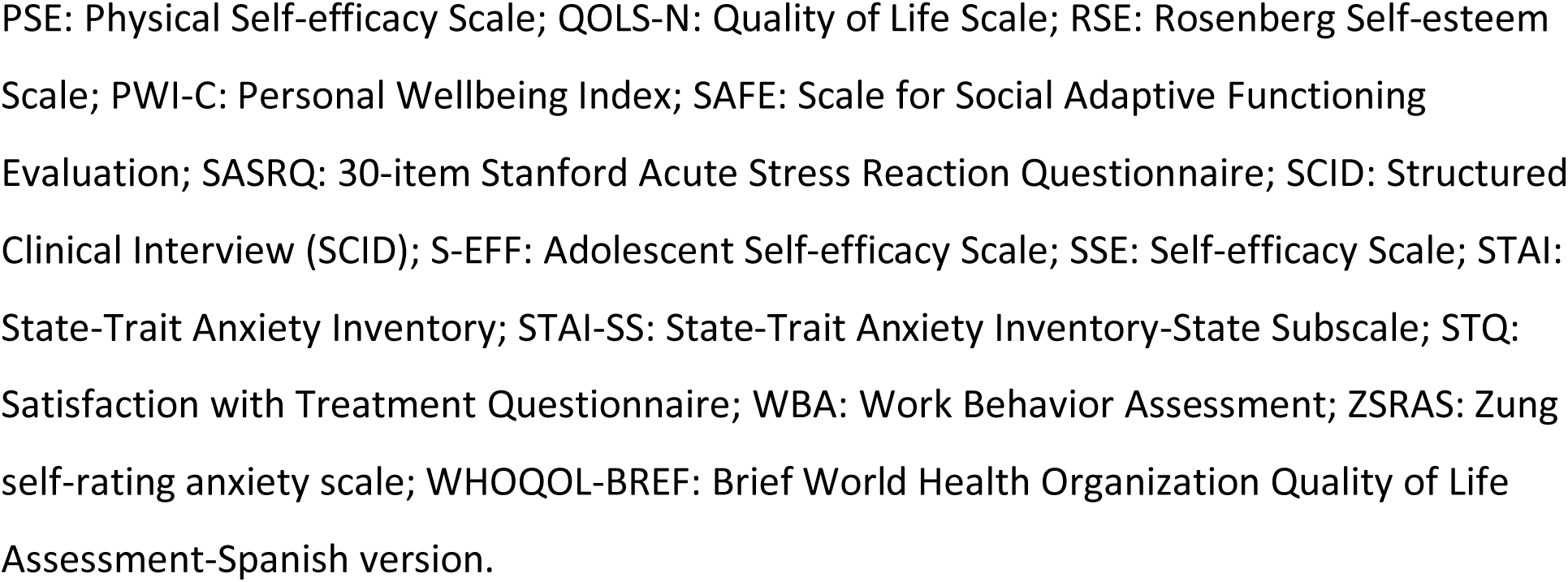
Study characteristics.

#### 3.3.1 Independent Checks

Two authors independently assessed RCTs identified from the search for inclusion first based on information included in the title and abstract and then by the retrieval of the full-text article. Any confusion or clarity around studies was resolved with one of the co-authors.

#### 3.4 Data Analysis and Synthesis

Percentages were calculated on CONSORT 2010 data to assess the quality of evidence. These descriptive information are presented in the findings subsection, Quality of Evidence. Furthermore, a qualitative narration was provided to describe the effectiveness of randomised control trials on ecotherapy interventions for the treatment of psychiatric disorders (see *Findings* subsection *Reported Effects of Ecotherapies)*

## 4. Findings

### 4.1 Description of Included Studies

Eighteen studies were included in the review. Eight studies included participants with a diagnosis of depression (Antonioli &Reveley, 2005; Berman, et al. 2012; Hoffmann et al., 2009; Kim, et al. 2009; McCaffrey, 2007; Pedersen et al., 2012; Shin et al., 2012), two included patients with both anxiety and depression (Berget et al., 2011; Le Roux &Kemp, 2009), one included patients with depression and comorbid obesity (Irandoust &Taheri, 2017), two included patients with substance dependence disorder (Wesley et al., 2009; Wesley et al., 2006), three included patients with schizophrenia (Barak et al., 2001; Villalta et al., 2009; Zhu, et al. 2016), one included patients with depression, schizophrenia or bipolar disorder (Kam &Siu, 2010), one included patients with schizophrenia, affective disorders, anxiety and personality disorders (Berget et al., 2008), and one included patients with post-traumatic stress disorder (Gelkopf et al., 2013). A total of 1,025 participants (N=17) between the age range of 18 - 75 years were included across thirteen RCT studies. The remaining five RCTs did not provide information on age (reference the five studies here). Ninety participants dropped out of the trials due to any cause (N=13). The participants were recruited as out-patient (N= #?) or through in-patient care facilities (e.g. rehabilitation centre, inpatient psychiatric care, hospitals and senior centres; N = #?). The eighteen studies (duration: 3 sessions to one year) (Hoffmann et al., 2009; Barak et al., 2001, respectively) included eight environmental therapy (i.e. gardening, forest therapy, horticultural therapy, nature adventure rehabilitation and adventure-based therapy) and ten animal-assisted therapy trials (i.e. dogs, horses or dolphins).

All eighteen studies had an intervention and comparison group. Eleven compared an environmental therapy or animal-assisted therapy to a control group (i.e. no treatment group) (Barak et al., 2001; Berget et al., 2011; Berget et al., 2008; Berman, 2012; Gelkopf et al., 2013; Hoffmann et al., 2009; Irandoust &Taheri, 2017; Le Roux &Kemp, 2009; Pedersen et al., 2012; Shin et al., 2012; Wesley et al., 2009), whereas seven trials compared these interventions to an alternative intervention (Antonioli &Reveley, 2005; Kam &Siu, 2010; Kim et al., 2009; McCaffrey, 2007; Villalta et al., 2009; Wesley, 2006; Zhu, et al., 2016). The primary outcomes that were assessed include: treatment efficacy (on the CGI-I or similar), symptom severity, and/or dropout rates. Secondary outcome measures assessed included self-efficacy, perceived control, hope, quality of life, life skills, and coping.

Seven studies were supported by funding (Berget et al., 2011; Berget et al., 2008; Berman, 2012; McCaffrey, 2007; Pedersen et al., 2012; Villalta et al., 2009; Zhu, et al. 2016). Studies were conducted in several countries, namely: China, Germany, Honduras, Iran, Israel, Norway, South Africa, South Korea, Spain and the United States of America. Twelve studies are single-centre trials (Barak et al., 2001; Berman, 2012; Gelkopf et al., 2013; Hoffmann et al., 2009; Irandoust &Taheri, 2017; Kam &Siu, 2010; Kim, et al., 2009; Le Roux &Kemp, 2009; Shin et al., 2012; Villalta et al., 2009; Wesley et al., 2009; Zhu, et al., 2016) and five are multi-centre trials (Antonioli &Reveley, 2005; Berget et al., 2011; Berget et al., 2008; McCaffrey, 2007; Pedersen et al., 2012).

### 4.2 Assessment of Quality of Evidence

Each included RCT was assessed across 25 criteria (See *Table 2: CONSORT 2010 Data*). Forty-four percent of the RCTs described the method used to generate random allocation sequence (Barak et al., 2001; Berget et al., 2008; Gelkopf et al., 2013; Kam &Siu, 2010; Pedersen et al., 2012; Villalta et al., 2009; Berget et al., 2011; Zhu et al., 2016) and the type of randomisation used (for example, block randomization) (Antonioli &Reveley, 2005; Berget et al., 2008; Gelkopf et al., 2013; Kam &Siu, 2010; Pedersen et al., 2012; Wesley et al., 2009; Berget et al., 2011; Zhu, et al., 2016). Thirty-three percent of the trials provided the numbers of participants who were randomly assigned, received intended treatment, and were analysed for the primary outcome (Antonioli &Reveley, 2005; Barak et al., 2001; Berget et al., 2008; Kam &Siu, 2010; Kim, et al., 2009; Irandoust &Taheri, 2017). Eighty-three percent of the trials did not provide information on the mechanism used to implement the random allocation sequence and any steps taken to conceal the sequence until interventions were assigned (expect for Berget et al., 2008; Kam &Siu, 2010; and, Zhu, et al., 2016). The blinding of participants and personnel was difficult to assess given that the ‘nature’ interventions could not be blinded. Two trials reported that they were single blind (Antonioli &Reveley, 2005; Kam &Siu, 2010). Zhu et al. (2016) was the only trial to be identified as a double-blind study, however. For attrition bias, losses and exclusions after randomisation, together with reasons were only reported by three trials (Antonioli &Reveley, 2005; Barak et al., 2001; Berget et al., 2008). Sixty-two percent of the trials reported primary and secondary outcomes, for each group. However, only five trials reported the estimated effect size and its precision (Antonioli &Reveley, 2005; Kam &Siu, 2010; Kim, et. al., 2009; Wesley et al., 2009; Zhu, et al., 2016). Binary outcomes were reported for each group for three trials, however the presentation of both absolute and relative effect sizes was not reported (Berget et al., 2008; Kim, et. al., 2009; Pedersen et al., 2012). Trial limitations, addressing sources of potential bias, imprecision, and, if relevant, multiplicity of analyses were reported by thirty-nine percent of the included trials (Antonioli &Reveley, 2005; Berman, 2012; Kam &Siu, 2010; Kim, et al. 2009; Hoffmann et al., 2009; Irandoust &Taheri, 2017; Zhu, et al., 2016). Generalisability was addressed in forty four percent of the trials. However, the balancing of benefits and harms were only reported in thirty-three percent of studies. Fifty percent of the trials were funded and the protocols for each of these trials could not be found (Berget et al., 2011; Berget et al., 2008; Berman, 2012; McCaffrey, 2007; Pedersen et al., 2012; Villalta et al., 2009; Zhu, et al., 2016).

**Table 2:**
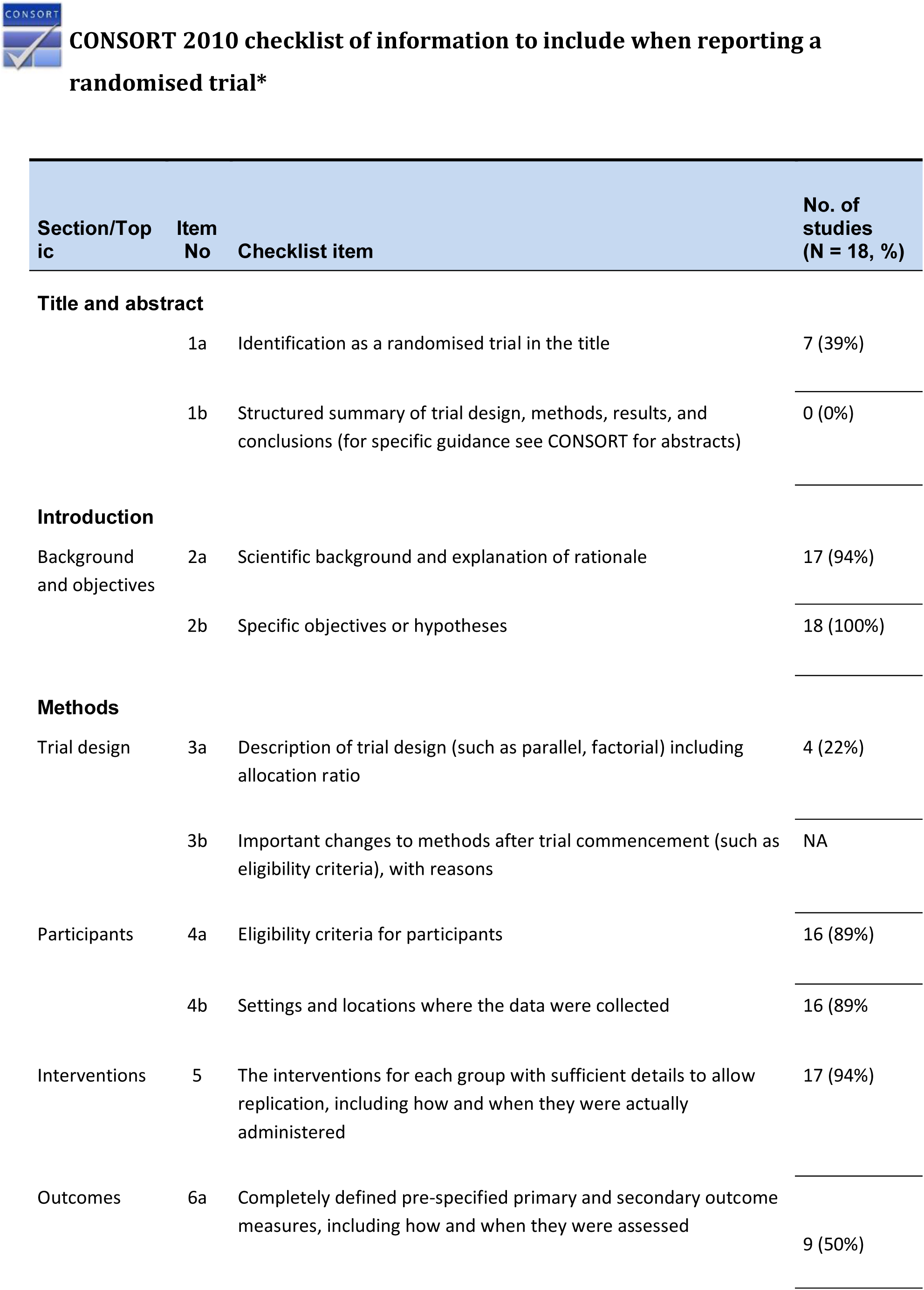
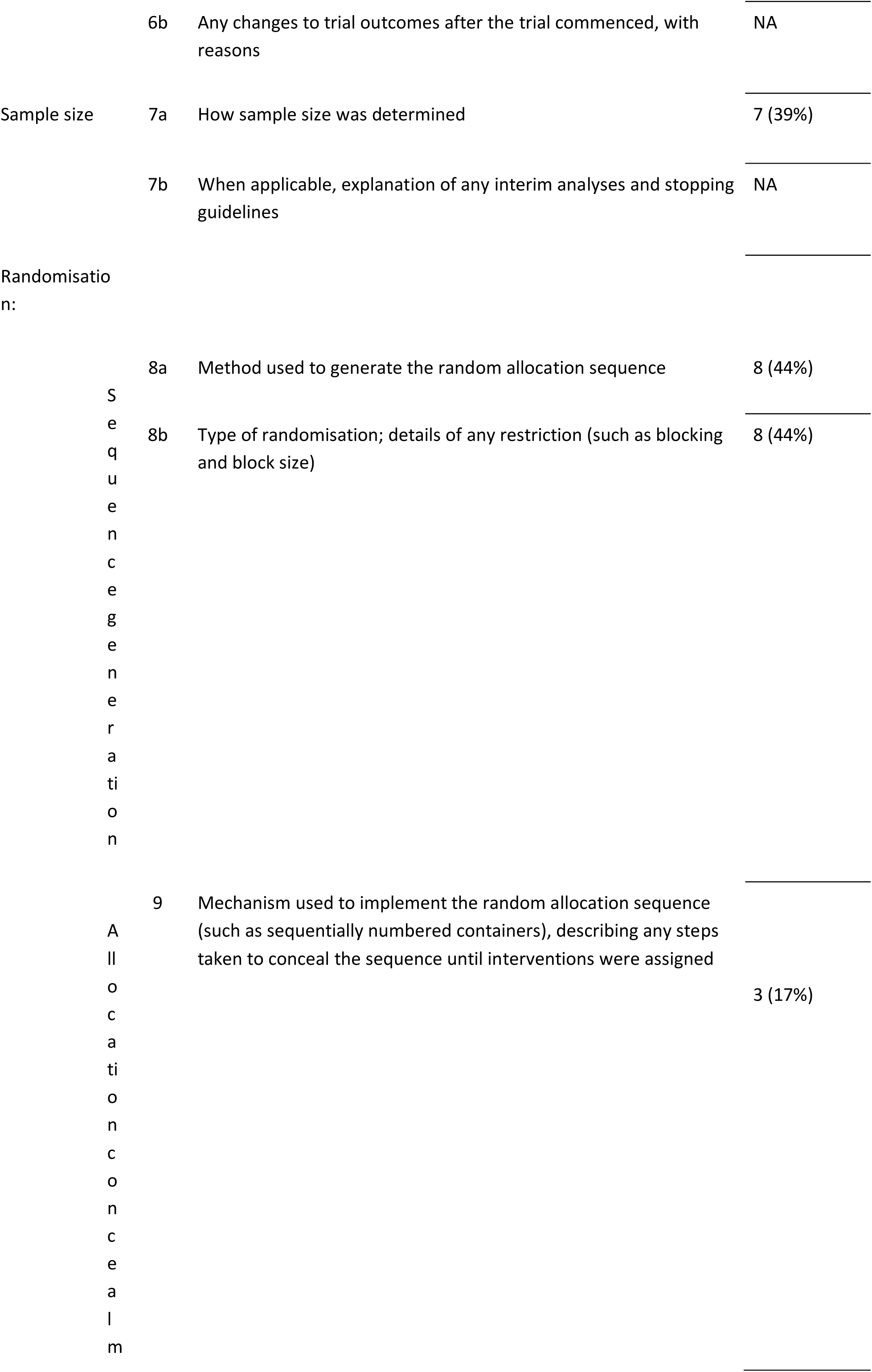
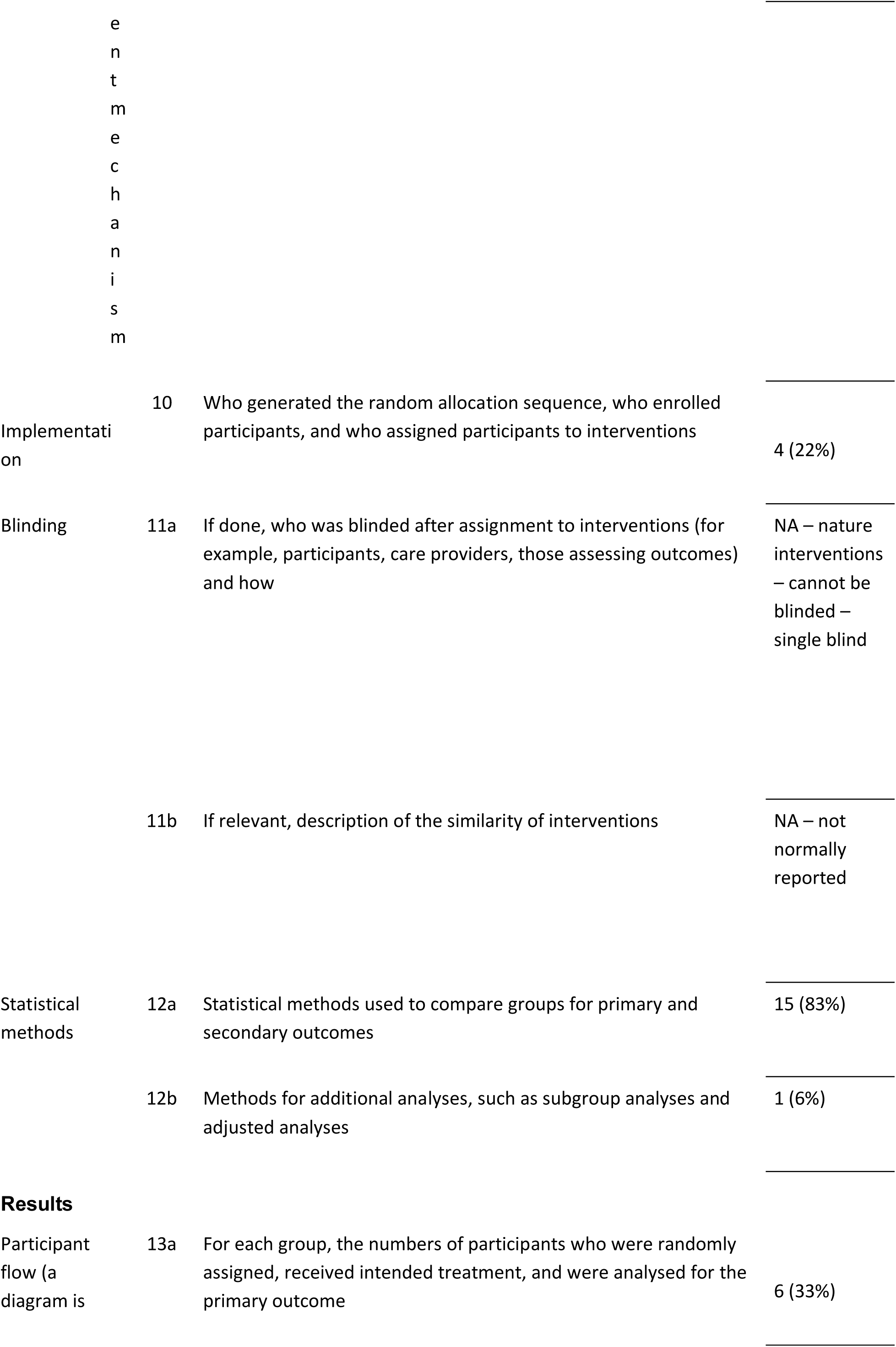
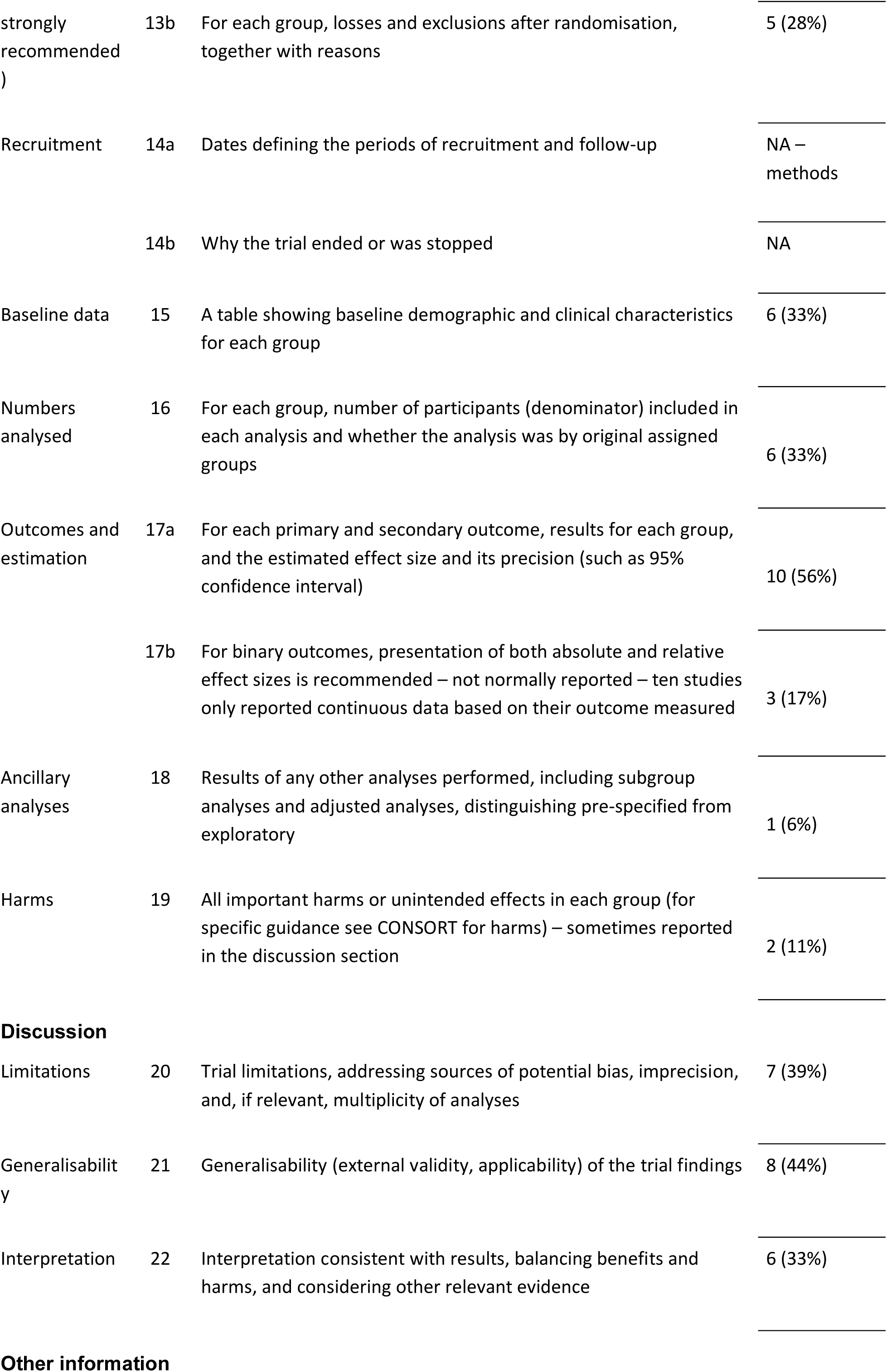
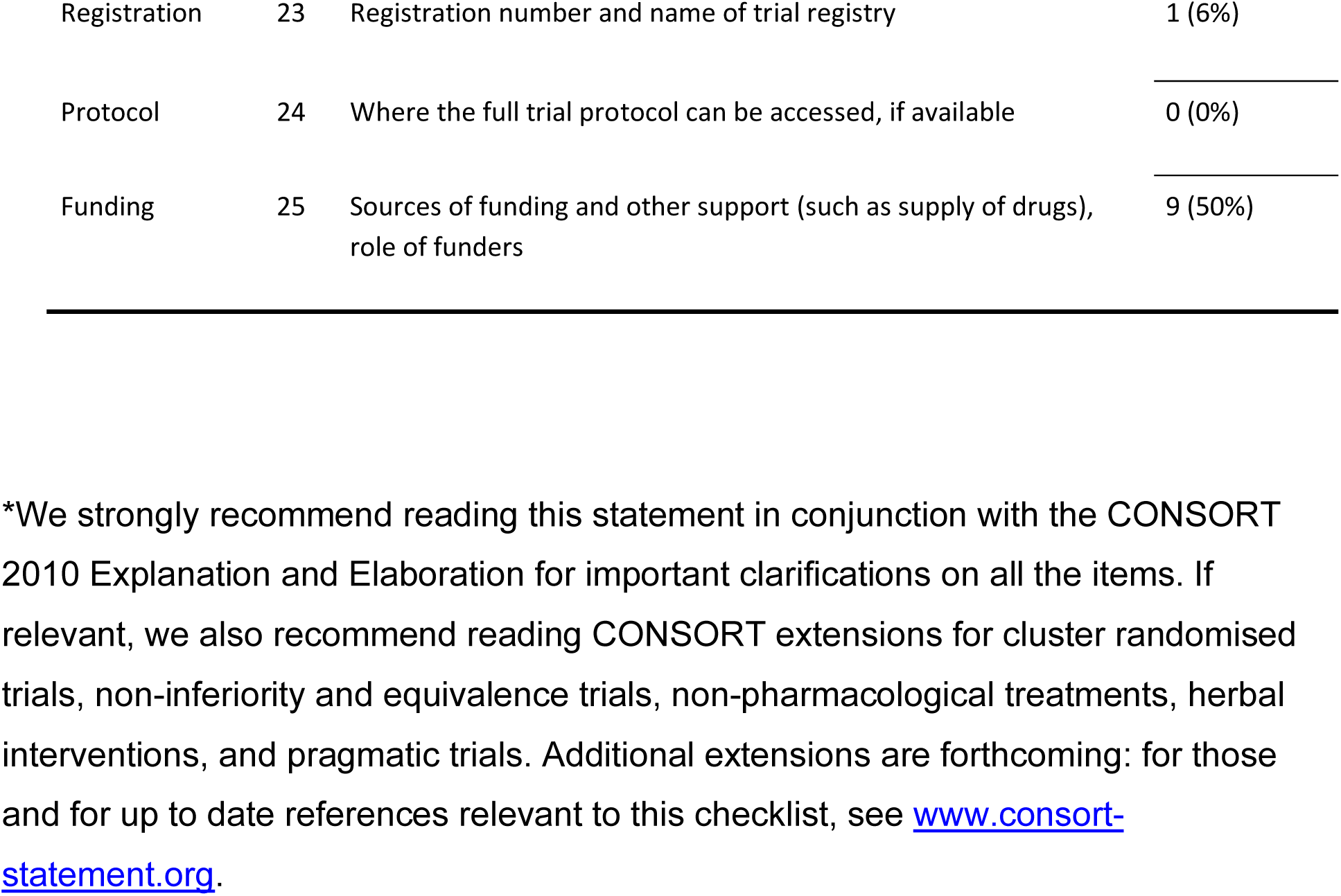
CONSORT 2010 Data.

### 4.3 Reported Effects of Ecotherapy

#### 4.3.1 Environmental Therapy

Eight studies used environmental interventions for the treatment of mental health disorders. Four of these trials focussed on depression, while one investigated depression and co-morbid alcohol use disorder. Berman, et al. (2012) found that individuals diagnoses with major depressive disorder showed significant cognitive improvements after nature walks than when compared to urban walks (p < .001). Nature walks also had a significant effect on positive affect when compared to urban walks (p < .001). However, changes in positive affects did not correlate with changes in cognitive improvements (ps > .19). The effect size of these improvements in a clinical population was said to be five times than that of a study previously conducted with a non-clinical population (Berman, et al. 2012; Berman, 2008).

Interventions were combined in some studies. McCaffrey (2007) found a significant difference for garden walks alone, garden walks with guided imagery, and/or art therapy for older adults with mild to moderate depression. In their study, group one, who received garden walks alone, reported that the natural environment gave them a sense of peace and serenity, as well as time for reflection about the meaning of life. The participants of group two, who received garden walks with a guided imagery leader, stated that crossing the bridge at the beginning of their walk was symbolic of leaving their problems of the past behind. Instead, they focused on a pleasant and fulfilling future. The additional group who painted in a natural environment found that it helped them with exploring who they were as an individual as well as the identification of their own strengths and weaknesses.

CBT-based psychotherapy for people living with depression that was applied in a forest environment significantly outperformed traditionally CBT-based psychotherapy (Kim, et al., 2009). The results suggested that CBT-based psychotherapy that is integrated into treatment in a forest environment can assist in the remission of depressive features. Additionally, Irandoust and Taheri (2017) studied the effects of vitamin D supplements alone and in combination outdoor exercise on depressive features in women with depression and comorbid obesity. The study found that a combination of outdoor physical activity and vitamin D supplements significantly improved depressive features (P = 0.001) compared to outdoor exercise or vitamin D alone. Furthermore, an improvement was observed amongst depressed patients with comorbid alcohol use disorder who attended a forest camp compared to the control group who continued to follow their daily routine. The difference between the two groups was statistically significant (p = 0.001). Participants who had substance use disorders receiving the intervention also found it less difficult to fall asleep (p = 0.001) and stay asleep (p = 0.009) (Shin et al., 2001).

Participants with psychiatric disorders (i.e. schizophrenia spectrum, major depressive and bipolar disorders) receiving horticultural therapy compared with those receiving a conventional workshop, reported a significant difference in change scores measured by a depression, anxiety and stress scale (p = 0.01). No significant differences in change scores measured by the personal well-being index (p ranges from 0 .08 to 0.79) and the work behaviour assessment (p = 0.84) were found, however. The participants also expressed that they enjoyed being in a natural environment and that this benefited them on a social, occupational and spiritual level (Kam, et al., 2010). In addition, Gelkopf (2013), found a significant reduction in post-traumatic symptoms (p = 0.05) when participants received a nature adventure rehabilitation (NAR) intervention compared with the waiting list control group. A reduction in depression symptoms (p = 0.05), emotional and social quality of life (p = 0.01), perceived control over illness (p = 0.01), and hope and functioning (p = 0.001) wasalso observed. Overtime these reductions in symptoms were significantly related to an improved sense of personal control.

One randomised, double-blind, placebo-controlled study investigated the treatment effect of horticultural therapy on people living with schizophrenia (Zhu, et al., 2016). Zhu et al., (2016) found that patients who were on antipsychotics and living with schizophrenia symptoms (positive and negative) significantly improved (p = 0.001) when exposed to horticultural therapy compared to the placebo-control group.

### 4.3.2 Animal-Assisted Therapy

Ten animal-assisted therapy trials were found. Participants with a diagnosis of depression reported reduced symptoms after two weeks of exposure with animal-assisted therapy with dolphins than the control outdoor nature programme group they were exposed to. These findings were observed on the Hamilton Depression Rating Scale (p = 0.002) and the Beck Depression Inventory (P = 0.006) (Antonioli &Reveley, 2005). Hoffmann et al. (2009) found a significant reduction in hospitalized patients with major depression and state anxiety, after the presence of a dog in the room (p = 0.016) in their post-treatment controlled crossover trial.

Villalta-Gil et al., (2012) found that there were no significant differences in the treatment of schizophrenia for inpatients in the treatment group who received dog-assisted psychotherapy and the control group who received psychotherapy alone. Nevertheless, those receiving dog-assisted therapy had significant improvements with social contact (p = 0.041), positive (p = 0.005) and negative symptom dimensions (p = 0.005), and quality of life related with social relationships (p = 0.024) when compared to those receiving treatment as usual.

A 12-week study assessing farm animals (i.e. cows, horses, and goats) and their effect on depression among psychiatric patients reported that anxiety and depression symptoms did not decrease during the intervention, although these symptoms were significantly lower at follow-up (p = 0.002) compared to the treatment-as-usual control group. Patients with the largest reduction in depression symptoms also experienced an increase in coping (p = 0.0006), mood (p = 0.049), self-esteem (p = 0.004), and extroversion (p = 0.06). No significant correlations were found with working ability or physical contact with animals, although physical contact with farm animals was strongly correlated with improved mood (p < 0.0001) (Berget et al., 2011). Furthermore, residents in a long-term care facility also report a significant difference in the reduction of depression symptoms (p = 0.017) when exposed to animal-assisted activities compared to those receiving treatment as usual. There was however no evidence of a difference in the reduction of anxiety symptoms for both groups (Le Roux 2009). Pedersen (2012) examined the effect of a 12-week farm animal-assisted therapy intervention on levels of depression, anxiety, and self-efficacy in people with clinical depression. A significant decline in symptoms of depression (p = 0.003) was found in the intervention group compared to the wait-list control group at endpoint. A significant increase in self-efficacy scores (p = 0.045) was also reported. However, no significant decrease in state anxiety symptoms was found (p = 0.059).

Amongst patients diagnosed with schizophrenia, the use of animal-assisted therapy did not demonstrate statistically significant improvements of instrumental and self-care, but improvements in social functioning were statistically significant compared to treatment as usual (Barak et al., 2001). Furthermore, a significant increase in self-efficacy was reported for psychiatric patients who were exposed to animal-assisted therapy with farm animals after six months of follow-up compared to treatment as usual (Berget et al., 2008). Although, no changes in quality of life were reported. Coping ability did show a positive change in scores for the treatment group. Nonetheless, patients with the largest increase in self-efficacy during the intervention also showed the largest increase in coping (p = 0.0029). Similarly, the patients with the largest increase in coping strategy reported the largest improvement in mood (p = 0.02), and they favoured to a larger extent physical contact with the animals (p = 0.03).

Finally, two trials investigated substance dependence and animal-assisted therapy.Wesley et al. (2006) and Wesley et al. (2009) reported that the therapeutic alliance was enhanced with the addition of a therapy dog. The addition of a therapy dog also lowered heart and diastolic blood pressure rates than those without exposure to a therapy dog. These results were not found amongst patients with a dual diagnosis.

## 5. Discussion

We found eighteen randomised control trials utilising an ecotherapy interventions (i.e. eight environmental therapy interventions and ten animal-assisted therapy interventions). A significant reduction in depression symptoms was reported by eleven RCTs (Antonioli &Reveley, 2005; Berman, et al., 2012; Berget et al., 2011; Gelkopf et al., 2013; Irandoust &Taheri 2017; Kam &Siu, 2010; Kim, et al., 2009; Le Roux &Kemp, 2009; McCaffrey, 2007; Pedersen et al., 2012; Shin et al., 2012), while a number of studies also reported a significant reduction in anxiety (Kam &Siu, 2010; Hoffmann et al., 2009; Le Roux &Kemp, 2009; Pedersen et al., 2012), post-traumatic stress symptoms (Gelkopf et al., 2013), schizophrenia (Barak et al., 2001; Zhu, et al., 2016) and addictive disorders (Wesley et al., 2009; Wesley et al., 2006). The evidence of this study on ecotherapy interventions for people living with mental health conditions is consistent with findings of previous systematic reviews on environmental and animal-assisted therapies in healthy populations. For instance, a significant reduction in depressive and anxiety features were found in systematic reviews on environmental therapies by Wen, et al. (2019) and Lee, et al. (2017).

The study has a number of limitations. It only searched articles published in the English language and underwent two updates since the primary systematic review. A meta-analysis was not performed owing to the heterogeneity of evidence.

There are also limitations to the evidence of these studies. The findings of this review specifically look at mood, anxiety, substance dependence, schizophrenia, and stressor-related psychiatric disorders. The studies and sample size of the included studies are few and small and this raises concerns around the overestimation of the effects of these interventions. There were more single-centre than multi-centre trials which may have biased the study findings and its generalisability across institutions (Higgins &Greens, 2011). Studies specifically assessing depression and anxiety used self-report measures. The lack of blinding and the reporting of effect estimates may have also biased the findings. Furthermore, findings from the assessment of quality of evidence suggest inconsistencies in standards of reporting across RCTs. These limitations of evidence are similar to those mentioned by other systematic reviews conducted on ecotherapies in healthy populations (Kamioka, et al., 2014; Kondo, et al., 2018; Wen, et al., 2019; Roberts, et al., 2019).Considering the complexity and multiple confounding variables, the appropriateness of RCTs for ecotherapy modalities is in question and should be further explored (Gabrielsen, 2016).

## 6. Conclusion

There is suggestive, but not conclusive, evidence for the efficacy of some ecotherapy interventions in mental disorders. Environmental therapies were effective in both the improvement and treatment of depressive, post-traumatic stress, alcohol use and schizophrenia spectrum disorders. Animal-assisted therapies have been shown to be effective for depressive, anxiety, alcohol use and schizophrenia spectrum disorders. In particular, there are: four environmental therapy trials and five animal-assisted therapy trails for depression and/or anxiety; one environmental therapy trial for post-traumatic stress disorder; three environmental therapy and animal assisted therapy trials for general psychiatric disorders; one environmental therapy trial and two animal assisted therapy trials for substance use disorders; and, one environmental therapy and two animal-assisted trials for people living with schizophrenia spectrum disorder. The use of standard reporting guidelines may improve evidence quality of future ecotherapy RCTs, and provide a foundation meta-analysis of the evidence.

## Data Availability

Included in content of tables

